# SARS-CoV-2 Omicron: evasion of potent humoral responses and resistance to clinical immunotherapeutics relative to viral variants of concern

**DOI:** 10.1101/2021.12.14.21267772

**Authors:** Anupriya Aggarwal, Alberto Ospina Stella, Gregory Walker, Anouschka Akerman, Vanessa Milogiannakis, Fabienne Brilot, Supavadee Amatayakul-Chantler, Nathan Roth, Germano Coppola, Peter Schofield, Jennifer Jackson, Jake Y. Henry, Ohan Mazigi, David Langley, Yonghui Lu, Charles Forster, Samantha McAllery, Vennila Mathivanan, Christina Fichter, Alexandra Carey Hoppe, Mee Ling Munier, Hans-Martin Jack, Deborah Cromer, David Darley, Gail Matthews, Daniel Christ, David Khoury, Miles Davenport, William Rawlinson, Anthony D. Kelleher, Stuart Turville

## Abstract

Genetically distinct viral variants of severe acute respiratory syndrome coronavirus 2 (SARS-CoV-2) have been recorded since January 2020. Over this time global vaccine programs have been introduced, contributing to lowered COVID-19 hospitalisation and mortality rates, particularly in the first world. In late 2021, the Omicron (B.1.1.529) virus variant emerged, with significant genetic differences and clinical effects from other variants of concern (VOC). This variant demonstrated higher numbers of polymorphisms in the gene encoding the Spike (S) protein, and there has been displacement of the dominant Delta variant. We assessed the impact of Omicron infection on the ability of: serum from vaccinated and / or previously infected individuals; concentrated human IgG from plasma donors, and licensed monoclonal antibody therapies to neutralise virus in vitro. There was a 17 to 22-fold reduction in neutralisation titres across all donors who had a detectable neutralising antibody titre to the Omicron variant. Concentrated pooled human IgG from convalescent and vaccinated donors had greater breadth of neutralisation, although the potency was still reduced 16-fold. Of all therapeutic antibodies tested, significant neutralisation of the Omicron variant was only observed for Sotrovimab, with other monoclonal antibodies unable to neutralise Omicron in vitro. These results have implications for ongoing therapy of individuals infected with the Omicron variant.

## Introduction

At the beginning of November 2021, the VOC Delta represented over 98% of SARS-CoV-2 infections sequenced worldwide. Omicron was first identified in South Africa on November 9^th^, 2021 and has primarily driven the fourth viral wave of infection in South Africa. The gene encoding the Spike (S) protein of the Omicron variant typically has 30 amino acid polymorphisms, three deletions and one insertion. Whilst the number of substitutions far exceeds that of other VOCs, the number of changes alone is not an effective surrogate to predict virus fitness in relation to transmission or ability to evade a humoral response generated through prior infection and/or vaccination. From the study of current VOCs and other variants, Omicron unfortunately has key critical changes in Spike. These have three main functional effects. Firstly, Omicron has multiple changes within the receptor-binding domain (RBD) associated with antibody evasion and also Angiotensin Converting Enzyme (ACE)-2 binding affinity. This includes previously described polymorphisms known for their ability to evade antibodies, including those at positions 484 and 477 ^1^. Secondly, many changes in the RBD are associated with potential significant increases in angiotensin-converting enzyme 2 (ACE2) affinity. S477N, Q498R and N501Y are located at the extremes of the RBD–ACE2 interface and act to stabilise this interaction ^2^. Finally, Omicron shares changes at (P681H) and around (H655Yand N679K) the S1/S2 furin cleavage site. These polymorphisms are associated with increased S1/S2 furin cleavage and more efficient fusogenic entry mediated by the serine protease TMPRSS2 ^3^. Of note, the two previous VOCs that dominated in 2021, Alpha and Delta, both shared a change at position 681 at the Spike furin cleavage site.

Herein we used a rapid and sensitive platform that was developed specifically for the isolation and characterisation of SARS-CoV-2 variants with respect to their relative transmission threat in previously infected and vaccinated populations. Within a week of obtaining the first positive Omicron nasopharyngeal swab sample in Australia, we documented greater resistance to neutralisation of Omicron compared to VOC Beta, Gamma and Delta across serum from both ChAdOx1 nCoV-19 and BNT162b2 double dose vaccinated donors. Further quantification of immune evasion by Omicron was established using three complementary approaches. The first was using individual serum samples from patients recruited to ADAPT, a community-based cohort of approximately 200 patients followed from the time of diagnosis during all three waves of infection in Australia. We studied a set of 50 samples with the highest humoral responses during convalescence following early viral clade infections in 2020: Clade A and B in March 2020 and Clade 20F in August 2020 and then Delta from June 2021. In addition to the study of the convalescent patients, we tested the highest humoral responders following vaccination during convalescence (Figure 1A-E). In each setting, the peak responses were tested across all relevant VOCs, Beta, Gamma, Delta, and Omicron. We then tested five polyclonal human hyperimmune IgG batches that constitute pools of thousands of primarily US plasma donors collected around the peak of US vaccination (Figure 1F). This latter analysis establishes the extent of immune evasion at the population level, as the IgG is comprised of all plasma donors irrespective if they are convalescent and/or vaccinated. Finally, we tested the therapeutic monoclonal antibodies including sotrovimab, casirivimab, imdevimab, bamlanivimab, cilgavimab and tixagevimab.

**Figure 1.**
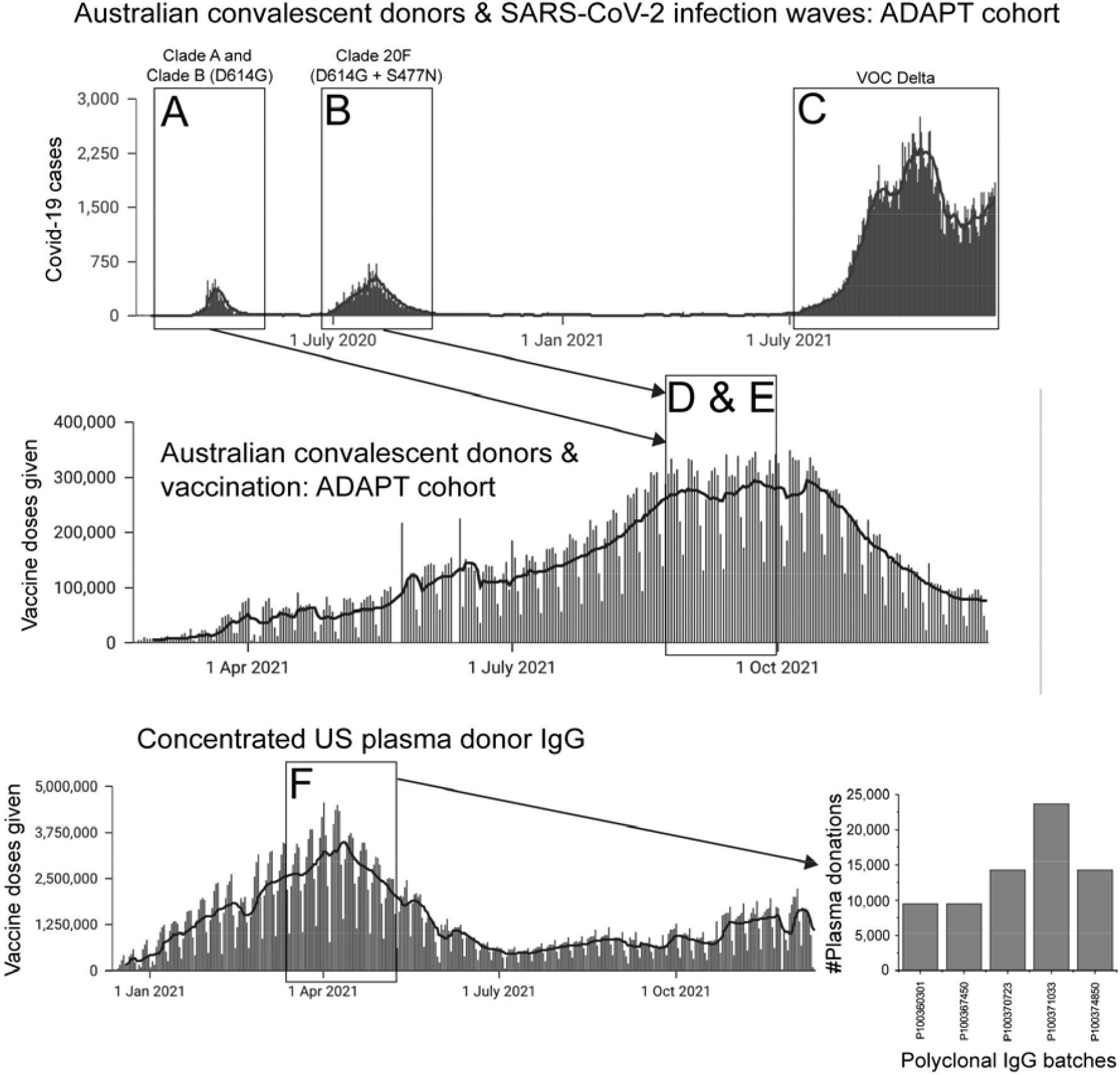
Source of high titre serology samples and polyclonal IgG obtained during the Covid-19 pandemic. Upper panel **A-C**: The ADAPT cohort and a summary of Covid-19 incidence over time in Australia. In brief, ADAPT is a community cohort of approximately 200 patients. The samples studied were from those with the highest neutralisation capacities to the ancestral strain. Herein we present the timeline of the Covid-19 pandemic illustrating the total number of daily Covid-19 cases Australia-wide during each wave of infection. **(A-C)** Convalescent serum donors from the Australian ADAPT cohort collected from **(A)** Clade A and Clade B (D614G) (first wave), **(B)** Clade 20F (D614G + S477N) (wave 2) and **(C)** the third (VOC Delta) wave. **(D-E)** Double dose vaccinated serum donors from the Australian ADAPT cohort collected during convalescence from **(D)** Clade A and Clade B (D614G) (first wave) and **(E)** Clade 20F (D614G + S477N) (wave 2). Vaccinated donors are split equally between ChAdOx1 nCoV-19 and BNT162b2 and represent the peak responses following vaccination (one month post second dose). **(F)** Concentrated human hyperimmune IgG from US plasma donors collected between March and May 2021 (left panel) including the number of plasma donations per polyclonal IgG batch tested (right panel). Not shown is the convalescent only polyclonal IgG, which was derived from US convalescent donors over the period of September and October 2020. This polyclonal IgG constituted approximately 5000 plasma donations over this period.

## Results and discussion

### Humoral evasion for Omicron relative to VOCs Beta, Gamma and Delta

The ability of Omicron and other VOCs (Beta, Gamma and Delta) to evade neutralising antibody responses was assessed using our rapid 20-hour live virus neutralisation platform (R-20) (manuscript submitted). Initial tests on Omicron using peak serological responses (one month post second dose) following two doses of BNT162b or ChAdOx1 nCoV-19 vaccines did not produce a recordable titre (Supplementary Fig. 1 and Supplementary table S1) across the 14 donors tested. To determine titre reductions, we selected high neutralisation titre serum samples from the Australian ADAPT cohort. Neutralisations performed with convalescent sera from early clade infections (Figure 2A and B; Supplementary Figure 2A and 2B) showed an average of 5.0-fold reduction for Beta, 1.6-fold for Delta and 2.1-fold for Gamma relative to the ancestral strain (A.2.2). All responses observed with Omicron were below the limit of detection of our assay (serum dilution 1:20). For convalescent sera obtained from Delta wave infections (Figure 2C; Supplementary Figure 2C) we observed a mean ID50 of 41.9 for Omicron compared with mean ID50 values of 770.5 for the ancestral strain, 211.5 for Beta, 317.5 for Gamma and 556.5 for Delta (Supplementary Table S1). Compared to the ancestral strain this resulted in a 20.7-fold reduction in neutralisation with Omicron (p<0.0001) compared to 3.9-fold for Beta (p<0.0024), 1.6-fold for Delta (p>0.9999) and 2.9-fold for Gamma (p<0.0365).

**Figure 2.**
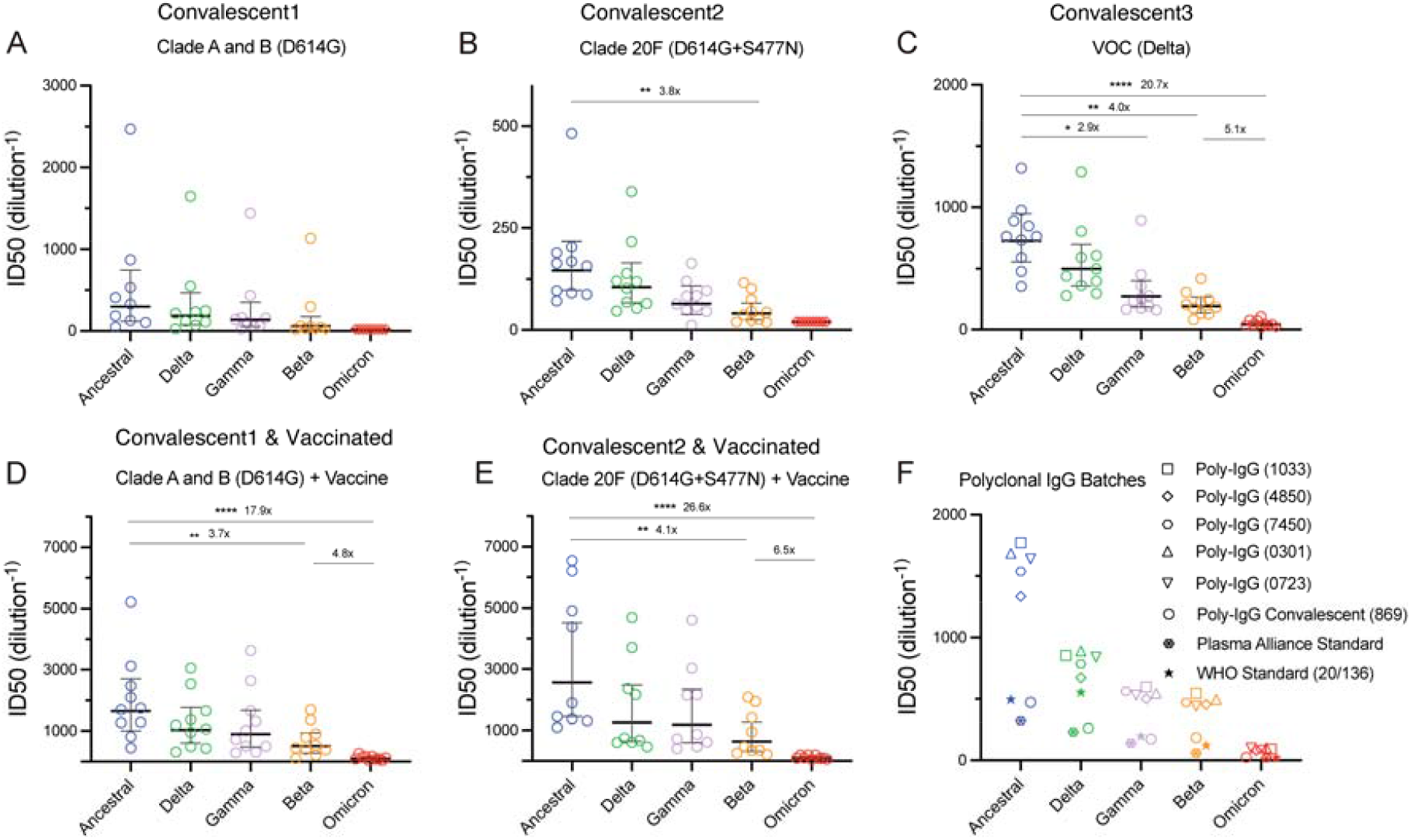
Humoral neutralisation of live SARS-CoV-2 variants in convalescent and vaccinated donors and with concentrated human IgG plasma samples. Neutralisation assays were performed in a high-throughput format in HAT-24 cells using live virus isolates from the variants of concern Delta (B.1.617.2), Gamma (P.1), Beta (B.1.351), Omicron (B.1.1.529) and the ancestral Wuhan-like virus with the original D614 background (A2.2) as a control. ID50 neutralisation titres are presented for five variants for convalescent donors from **(A)** Clade A and B (D614G) (first wave), **(B)** Clade 20F (D614G + S477N) (second wave) and **(C)** third (VOC Delta) wave. ID50 neutralisation titres presented for 5 live variants for vaccinated donors from **(D)** Clade A and B (D614G) (first wave) and **(E)** Clade 20F (D614G + S477N) (the second wave). **(F)** ID50 neutralisation titres across five live variants with concentrated polyclonal IgG from either convalescent and vaccinated donors or convalescent donors only. Data in **(A-F)** indicates the mean ID50 of technical replicates for individual samples (circles) with the geometric mean and 95% confidence interval shown for each variant. Titres below the limit of detection are indicated with red diamonds. Fold change reductions in ID50 neutralisation titres compare variants of concern to the ancestral variant, as well as Beta to Omicron. *p<0.05, **p<0.01, ***p<0.001, ****p<0.0001 for Kruskal Wallis test with Dunn’s multiple comparison test.

We next examined neutralisation responses in sera from convalescent donors from early clade infections who had subsequently been vaccinated with either the BNT162b or ChAdOx1 vaccine (Figure 2D and E and Supplementary Figure 2D and Supplementary Table S2; presented together as a single “vaccinated” group). While the vaccinated individuals showed a significant increase in neutralisation titres to the ancestral strain compared to convalescent donors (Supplementary Table S1), we again observed a 17.9 to 26.6-fold reduction in neutralisation against Omicron (p<0.0001) compared to 3.7 to 4.1-fold decrease observed for Beta (p<0.0028).

Similar results were obtained when we tested neutralisation responses against five polyclonal human IgG batches comprising of more than ten thousand pooled plasma donors collected during the peak of the US vaccine rollout (Figure 2F; Supplementary Figure 2F). There was a 16.8-fold reduction in neutralisation against Omicron (p<0.0001) compared with 3.3-fold decrease for Beta (p<0.0437). Similar fold reductions were also observed from polyclonal IgG that was collected from convalescent donors between September and October 2020 (fold reduction of 20.1-fold for Omicron versus 5.5-fold for Beta).

### Estimated fold reduction of Omicron and implications for vaccine efficacy

We next estimated the fold reduction in neutralisation for each variant within each cohort (with censoring). To do so, we grouped the data into three convalescent groups (Conv1-First wave; Conv2-Second wave; Conv3-Third wave (VOC Delta) see A, B and C in fig. 1), convalescent plus vaccinated, pooled polyclonal IgG and the WHO Standard G as a control. This data is summarised in Figure 3. Note that 3 cohorts had no detectable neutralisation against Omicron and thus were completely excluded from this aggregated analysis. For estimating the reduction in vaccine efficacy, Delta wave individuals were also excluded since they were exposed to the Delta Spike immunogen and may have a different cross-reactivity (though including this group changes the results only very slightly). The average fold reduction across all donors from the first wave including vaccinated individuals is 20.2 fold (95% CI: 16.7-24.3). Using this value, and the efficacy curve in Khoury et al ^4^ we estimated the efficacy and confidence intervals for BNT162b-vaccinated or boosted individuals (in the first few months after vaccination) (Table 1).

**Figure 3.**
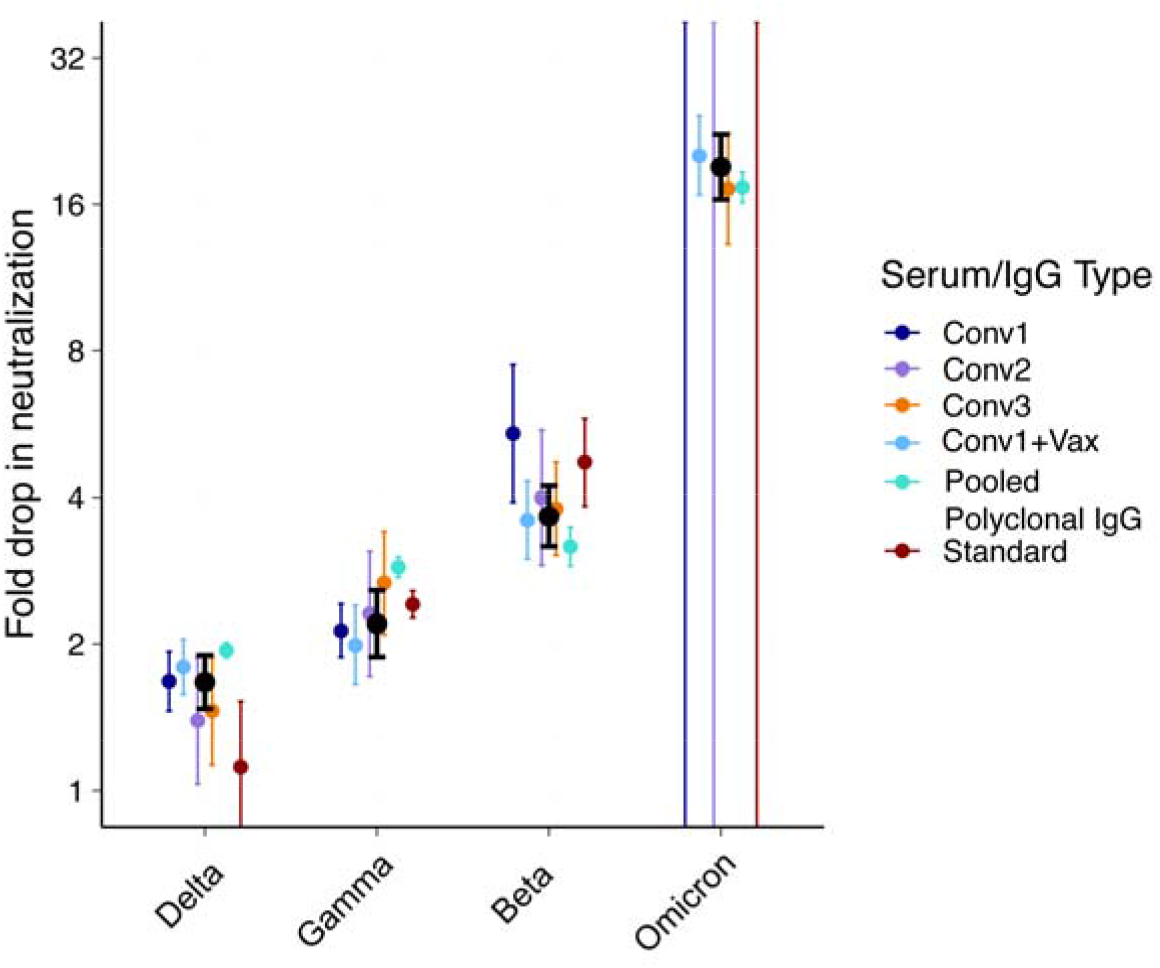
Summary of neutralisation reductions across Omicron and other variants of concern. Black dots represent the mean of all subjects (except for Omicron where only groups of individuals with detectable neutralisation against Omicron were included). Error bars are 95% CIs of the geometric mean. Unbounded error bars are present for groups where no detectable neutralisation was observed against Omicron.

**Table 1:** Estimates of Efficacy (95% CIs)

|  | With censoring |  |
| --- | --- | --- |
|  | BNT162b2 | mRNA boosted |
| Symptomatic | 37.2%<br>(22.2%-51.4%) | 74.4%<br>(60.9%-84.0%) |
| Severe* | 78.8%<br>(45.9%-93.6%) | 95.7%<br>(82.5%-99.1%) |

*\*Note that Vaccine Efficacy (VE) against severe outcomes is much less well validated as there is insufficient data available with which to parameterise the model at such low titres against severe outcomes, in addition to a lack of understanding of baseline severity with Omicron.*

### Modelling on vaccine efficacy based on Omicron fold evasion from humoral responses

In combining the data obtained from the 50 patients with the highest neutralisation titres in the ADAPT cohort with the population humoral snapshot using polyclonal IgG from pooled plasma donors, it is evident that the fold reduction in neutralisation of omicron similar. Given the starting neutralisation level of the average BNT162b2 vaccinee against the ancestral virus (2.4-fold of convalescent individuals) ^4^, a 20.2-fold drop brings the mean neutralisation level below the 50% protective level against symptomatic infection with Omicron. This neutralisation level corresponds to a predicted vaccine efficacy for otherwise naïve BNT162b2 vaccinated individuals of 37.2% (95% CI = 22.2-51.4%) against symptomatic infection with Omicron, and 74.4% (95% CI = 60.9%-84.0%) protection against severe infection (Table 1), in the first months after vaccination. Previously we have shown that mRNA vaccination of previously infected individuals produce neutralisation titres that are significantly higher than observed in current two-dose vaccination regimes ^5^. Thus, even with the 20.2-fold decrease in neutralisation titre, boosting with mRNA vaccines is predicted to provide significant protection from infection with Omicron (Table 1).

### Sotrovimab retains activity against Omicron

Bioequivalents of current therapuetic monoclonal antibodies (mAbs) were assessed for neutralisation, including sotrovimab, casirivimab, imdevimab, bamlanivimab, cilgavimab and Tixagevimab. Neutralisation by mAbs was assessed in VeroE6 cells, as we and others have previously found some mAbs to be unsuited for assessment in cell lines expressing human ACE2 ^6^. Sotrovimab (IC_50_= 1059 ng/mL) and Tixagevimab (IC_50_= 3490 ng/mL) neutralized the Omicron variant, although less effectively than the ancestral lineage. The reduction in Omicron IC50 potency was 2.8-fold and 73.8-fold for Sotrovimab and Tixagevimab, respectively. Casirivimab, Imdevimab, Bamlanivimab, Cilgavimab, and Ab-3467 did not neutralize at the highest concentrations tested (Table 2).

**Table 2:**
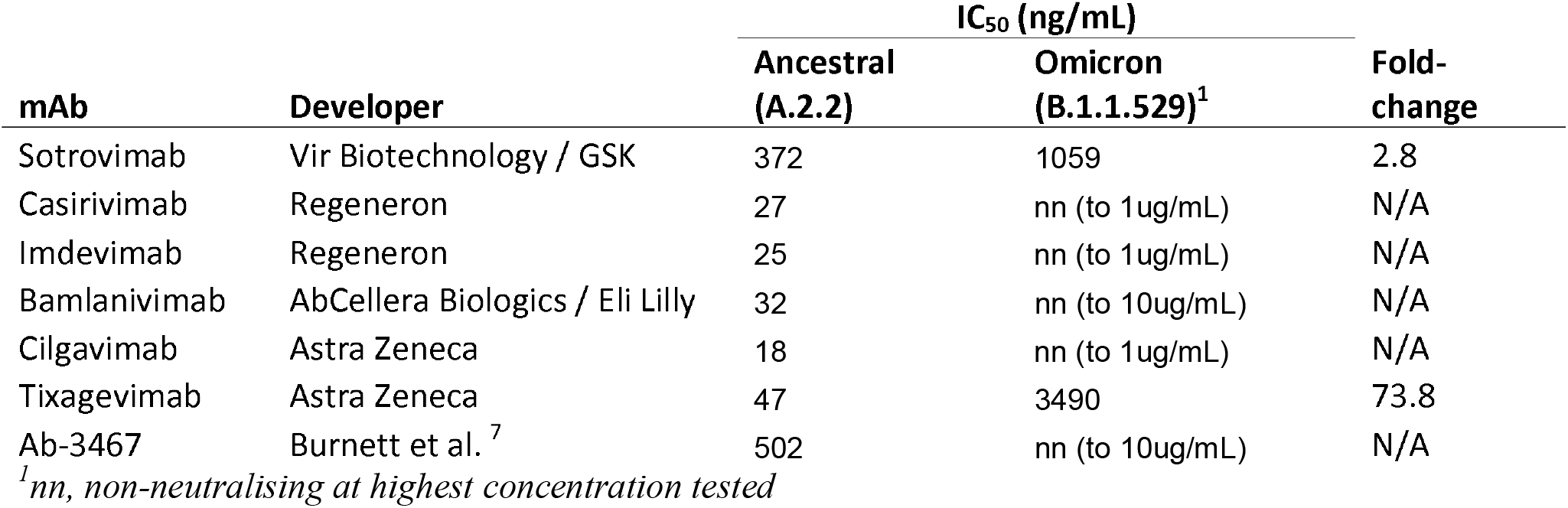
Neutralisation of SARS-CoV-2 omicron variant by commercially developed monoclonal antibodies and the class 4 Ab-3467.

| mAb | Developer | IC<sub>50</sub> (ng/mL) |  | Fold-change |
| --- | --- | --- | --- | --- |
|  |  | Ancestral (A.2.2) | Omicron (B.1.1.529)<sup>1</sup> |  |
| Sotrovimab | Vir Biotechnology / GSK | 372 | 1059 | 2.8 |
| Casirivimab | Regeneron | 27 | nn (to 1ug/mL) | N/A |
| Imdevimab | Regeneron | 25 | nn (to 1ug/mL) | N/A |
| Bamlanivimab | AbCellera Biologics / Eli Lilly | 32 | nn (to 10ug/mL) | N/A |
| Cilgavimab | Astra Zeneca | 18 | nn (to 1ug/mL) | N/A |
| Tixagevimab | Astra Zeneca | 47 | 3490 | 73.8 |
| Ab-3467 | Burnett et al. <sup>7</sup> | 502 | nn (to 10ug/mL) | N/A |

<sup>1</sup>nn, non-neutralising at highest concentration tested

The Omicron variant contains several mutations at RBD sites previously thought to be highly conserved that are the target of antibody therapeutics. For example, S371L, S373P, and S375F form part of the class 4 epitope, affecting previously described class 4 antibodies such as Ab-3467 that broadly neutralize sarbecoviruses ^7,8^. Additional mutations unique to Omicron at sites 417, 440, 446, and 493 are likely to contribute to the lack of the neutralisation of other therapeutic antibodies. The class 3 antibody Sotrovimab targets a highly conserved region of sarbecovirus RBD ^9^ and retains neutralising activity against Omicron in live virus neutralisation, with two mutations within its epitope (G339D & N440K) likely resulting in only a moderate reduction in potency observed against Omicron relative to the ancestral SARS-CoV-2 lineage. As new variants of SARS-CoV-2 emerge that have altered transmissibility and disease phenotype, the availability of therapeutic and prophylactic mAbs that remain broadly active is essential. Although the retention of neutralising activity by Sotrovimab against Omicron is promising, the complete loss of activity of many other monoclonals remains a concern, and the development of new and improved monoclonal antibody modalities is urgently warranted.

To conclude, Omicron represents a significant challenge to the existing two dose vaccination strategy presently adopted by many countries globally. Whilst the VOCs Beta and Gamma also represented challenges to vaccine efficacy, there are two defining features of Omicron that provide additional concerns. Firstly, as observed herein the fold evasion to humoral immunity is significantly greater with Omicron than all other VOCs. Secondly, unlike Beta and Gamma, Omicron is gaining momentum in global prevalence in areas where Delta has dominated in late 2021. Whilst boosters utilising the same Clade A Spike may increase antibody titres to Omicron, development of variant specific boosters may be more pragmatic in the longer term if Omicron persists. The latter will be very important in those groups that may have a limited titre, such as in the elderly or immunocompromised. Fortunately for the latter at risk groups, certain immunotherapeutic treatments like Sotrovimab appear to maintain potency and remain relevant for treatment in Omicron cases.

## Materials & Methods

### Human sera and ethics statement

The Adapting to Pandemic Threats (ADAPT) cohort is composed of RT-PCR–confirmed convalescent individuals (incl. some subsequently vaccinated) recruited in Australia since 2020 ^10^. The Adapting to Pandemic Threats (ADAPT) cohort is composed of RT-PCR– confirmed convalescent individuals (incl. some subsequently vaccinated) recruited in Australia since 2020 ^10^. Serum from healthy volunteers vaccinated with ChAdOx1 and BNT162b2 was collected 4 weeks post second-dose vaccination. A subgroup of ADAPT participants with the highest neutralization responses were selected for this study (Supplementary Table S2). All human serum samples were obtained with written informed consent from the participants (2020/ETH00964; 2020/ETH02068).

### Other immunoglobulin products

For monoclonal antibodies, DNA sequences encoding the variable domain sequences of the therapeutic monoclonal antibodies sotrovimab, casirivimab, imdevimab, bamlanivimab, cilgavimab and tixagevimab were generated by gene synthesis, cloned into human IgG1 expression vectors, and produced in CHO cells ^7^.

### Polyclonal Immunoglobulin preparations and anti-SARs-CoV-2 hyperimmune globulin

A CoVIg-19 Plasma Alliance (Poly IgG Convalescent 869) was formed in 2020 between major plasma pharmaceuticals including CSL, Takeda, Octapharma and Sanquin with an aim to develop a COVID-19 immunoglobulin therapy. As part of that initiative, CSL Behring manufactured anti-SARS-CoV-2 hyperimmune globulin (CoVIg). Approximately 5000 convalescent donor plasma units were collected between September and October 2020, exclusively from SARS-CoV2 convalescent donors after COVID-19 confirmation (Kober et al., 2021), the immunoglobulin purified using the licensed and fully validated immunoglobulin manufacturing process used for Privigen ^11^, notionally similar to others ^12^. Five IVIG lots (Poly IgG 1033, 4850, 7450, 0301, 0723) manufactured using the Privigen process described by Stucki et al. ^11^ included US plasma collected by plasmapheresis from a mixture of vaccinated with SARS-CoV-2 mRNA vaccines, convalescent and non-convalescent donors (source plasma, n between 9495-23,667 per batch) majority of donations collected between April and June 2021. The WHO international reference standard for SARS-CoV-2 neutralization (NIBSC 20/136) was obtained from ^13^.

### Cell culture

Hek293T cells stably expressing human ACE2 and TMPRSS2 were generated by lentiviral transductions as previously described ^10^. A highly permissive clone (HAT-24) was identified through clonal selection and used for this study. The HAT-24 line has been extensively cross-validated with the VeroE6 line (manuscript submitted). HAT-24 cells and VeroE6-TMPRSS2 (CellBank Australia, JCRB1819) were cultured in DMEM-10%FBS and VeroE6 cells (ATCC® CRL-1586™) in MEM-10%FBS. All cells were incubated at 37°C, 5% CO_**2**_ and >90% relative humidity.

### Viral isolation, propagation, and titration

All laboratory work involving infectious SARS-CoV-2 occurred under biosafety level 3 (BSL-3) conditions. Diagnostic respiratory specimens testing positive for SARS-CoV-2 (RT-qPCR, Seegene Allplex SARS-CoV-2) were sterile-filtered through 0.22 µm column-filters at 10,000x g and serially diluted (1:3) on HAT-24 cells (10^**4**^ cells/well in 96-well plates). Upon confirmation of cytopathic effect by light microscopy, 300μL pooled culture supernatant from infected wells (passage 1) were added to VeroE6 cells in a 6-well plate (0.5 × 10^**6**^ cells/well in 2mL) and incubated for 48h. The supernatant was cleared by centrifugation (2000 xg for 5 minutes), frozen at -80 °C (passage 2), then thawed and titrated to determine median tissue culture infective dose (TCID50) on VeroE6 or VeroE6-TMPRSS2 cells according to the Spearman-Karber method ^14^. Viral stocks used in this study correspond to passage 3 virus, which were generated by infecting VeroE6-TMPRSS2 cells at MOI=0.025 for all variants with the exception of Omicron. For the latter Omicron was expanded only in the VeroE6 line, as lower titers were observed when expanding virus using the VeroE6-TMPRSS2 line. For all variants, passage 3 expansions were on for 24h before collecting, clearing, and freezing the supernatant as above. This ensures a similar infectivity to particle ratio across all variants. Sequence identity and integrity were confirmed for both passage 1 and passage 3 virus via whole-genome viral sequencing using an amplicon-based Illumina sequencing approach, as previously described ^15^. For a list of the viral variants used in this study see Supplementary table S3. Passage 3 stocks were titrated by serial dilution (1:5) in DMEM-5%FBS, mixing with HAT-24 cells live-stained with 5% v/v nuclear dye (Invitrogen R37605) at 1.6×10^**4**^/well in 384-well plates, incubating for 20h, and determining whole-well nuclei counts with an InCell Analyzer high-content microscope and IN Carta analysis software (Cytiva, USA). Data was normalised to generate sigmoidal dose-response curves (average counts for mock-infected controls = 100%, and average counts for highest viral concentration = 0%) and median lethal dose (LD_**50**_) values were obtained with GraphPad Prism software.

### Rapid high-content SARS-CoV-2 microneutralization assay with HAT-24 cells (R20)

Human sera or monoclonal antibodies were serially diluted (1:2 series starting at 1:10) in DMEM-5%FBS and mixed in duplicate with an equal volume of SARS-CoV-2 virus solution standardised at 2xLD_**50**_. After 1h of virus–serum coincubation at 37°C, 40μL were added to an equal volume of nuclear-stained HAT-24 cells pre-plated in 384-well plates as above. Plates were incubated for 20h before enumerating nuclear counts with a high-content fluorescence microscopy system as indicated above. The % neutralization was calculated with the formula: %N = (D-(1-Q)) × 100/D as previously described ^10^. Briefly, “Q” is a well’s nuclei count divided by the average count for uninfected controls (defined as having 100% neutralization) and D = 1-Q for the average count of positive infection controls (defined as having 0% neutralization). Sigmoidal dose-response curves and ID_**50**_ values (reciprocal dilution at which 50% neutralization is achieved) were obtained with GraphPad Prism software. Neutralization assays with VeroE6 cells were performed exactly as described above excepting that; input virus solution was standardised at 1.25 × 10^**4**^ TCID50/mL, cells were seeded at 5×10^**3**^ cells/well in MEM-2%FBS (final MOI = 0.05), plates were incubated for 72h, and cells were stained with nuclear dye only 1h before imaging.

### Estimating vaccine efficacy against Omicron

Vaccine efficacy against Omicron was estimated using the approach and model previously reported^4 16^. Briefly, we estimate vaccine efficacy (VE) for a given vaccine/cohort against a given variant based on the (log_10_) geometric mean (GM) neutralisation titre, normalised to the GM neutralisation titre (against ancestral virus) of convalescent individuals (exposed to ancestral virus). This normalised (log_10_) GM titre is given by *µ*, and the VE for a given *µ* is predicted using the equation

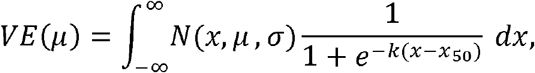

where, *N* is the probability density function of a normal distribution of the distribution in (log_10_) neutralisation titres with mean *µ* and standard deviation *σ*, and *x*_50_ and *k* are the associated with 50% protection and *k* determines the steepness of the logistic function. The parameters of a logistic function representing the (log_10_) normalised neutralisation titre parameters *σ = 0*.*46* (representing the spread of neutralisation titres in vaccinated individuals), *x*_50_ = log_10_ 0.20 and *k* = 3.1 were estimated previously by fitting vaccine efficacy data from randomised control trials across 7 SARS-CoV-2 vaccines^4^. Thus, vaccine efficacy of BNT162b2 vaccinated individuals against Omicron is estimated by estimating the normalised GM neutralisation titre of BNT162b2 vaccinated individuals against Omicron. Previously we have estimated that BNT162b2 vaccinated individuals have a GM neutralisation titre (against ancestral virus) that is 2.4-fold times the GM neutralisation titre of convalescent individuals (who were exposure to ancestral virus)^4^. Thus, to estimate the VE against Omicron of BNT162b2 vaccinated individuals, we calculate the normalised GM neutralisation titre of these individuals against Omicron. A 20.2-fold drop in neutralisation against Omicron is estimated to lead to a GM neutralisation titre in BNT162b2 vaccinated individuals against Omicron of 2.4/20.2 = 0.119-fold of the GM neutralisation titre of convalescent individuals (after exposure to ancestral virus) against ancestral virus (thus, *µ* = log_10_ 0.119 in the above equation to estimate vaccine efficacy for BNT162b2 vaccinated individuals against Omicron). Similar, we have previously shown that mRNA vaccines in previously infected individuals produce GM neutralisation titres that are approximately 12.0-fold greater than the GM titre of convalescent individual who were exposed to ancestral virus^5^. Thus, the 20.2-fold loss of neutralisation to Omicron is estimated to give a GM neutralisation titre against Omicron that is 0.594-fold of the GM of convalescent subjects against ancestral virus (after exposure to ancestral virus). Thus, to estimate vaccine efficacy for previously infected and mRNA vaccinated individuals we use *µ* = log_10_ 0.594 in the above equation. Confidence intervals for predicted vaccine efficacies were estimated using bootstrapping.

## Data Availability

All data produced in the present work are contained in the manuscript

## Supplementary Figures

**Supplementary Figure 1.**
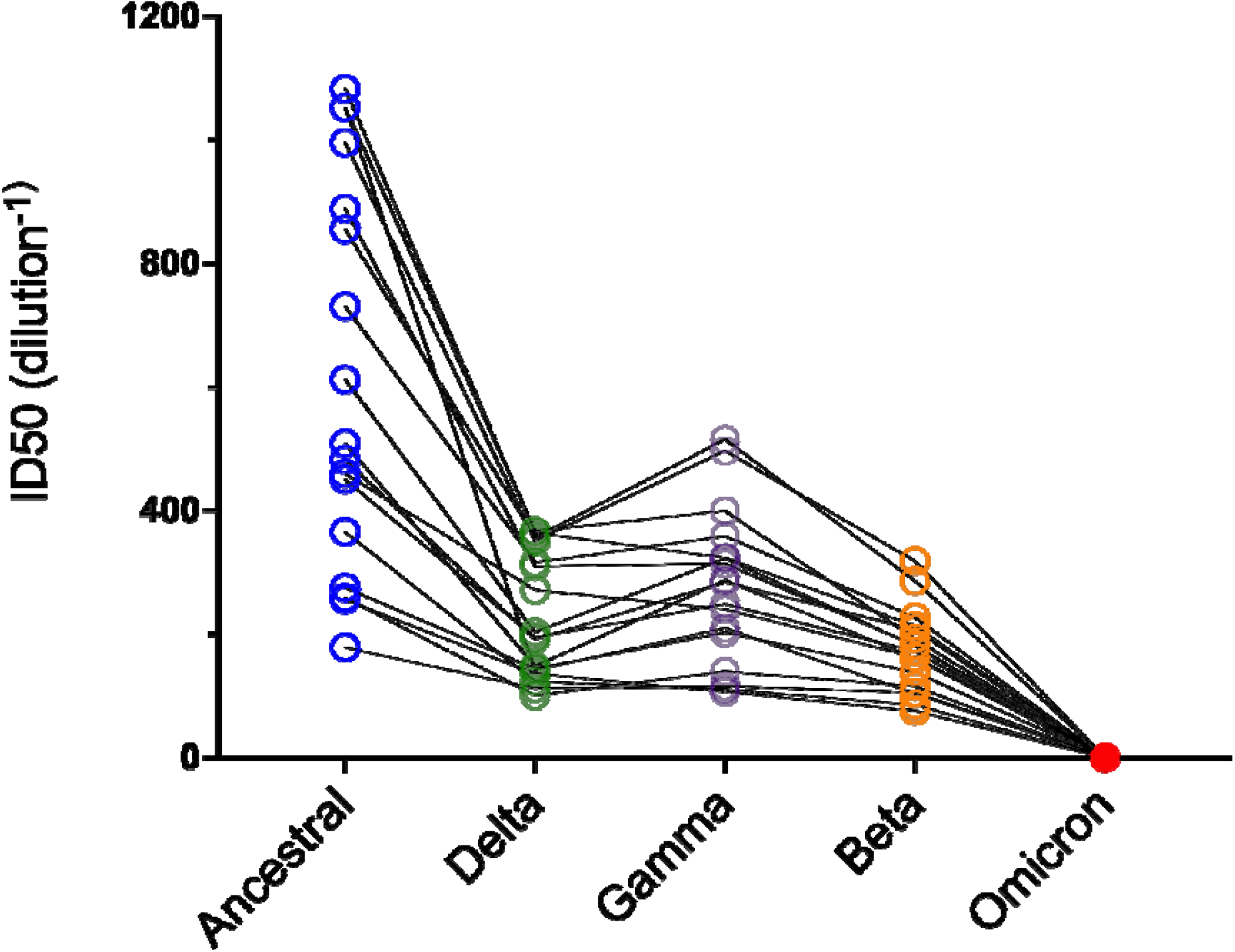
Initial humoral neutralisation of variant of concern Omicron in BNT162b or ChAdOx1-vaccinated individuals. Neutralisation assays were performed in a high-throughput format in HAT-24 cells using live virus isolates from the variants of concern; Delta, (B.1.617.2), Gamma (P1), Beta (B.1.351), Omicron (B.1.1.529) and the ancestral Wuhan-like virus with the original D614 background (A2.2) as a control. ID50 neutralisation titres across 3 live variants with BNT162b or ChAdOx1-vaccinated individuals. Data represents the mean of technical replicates in duplicate.

**Supplementary Figure 2.**
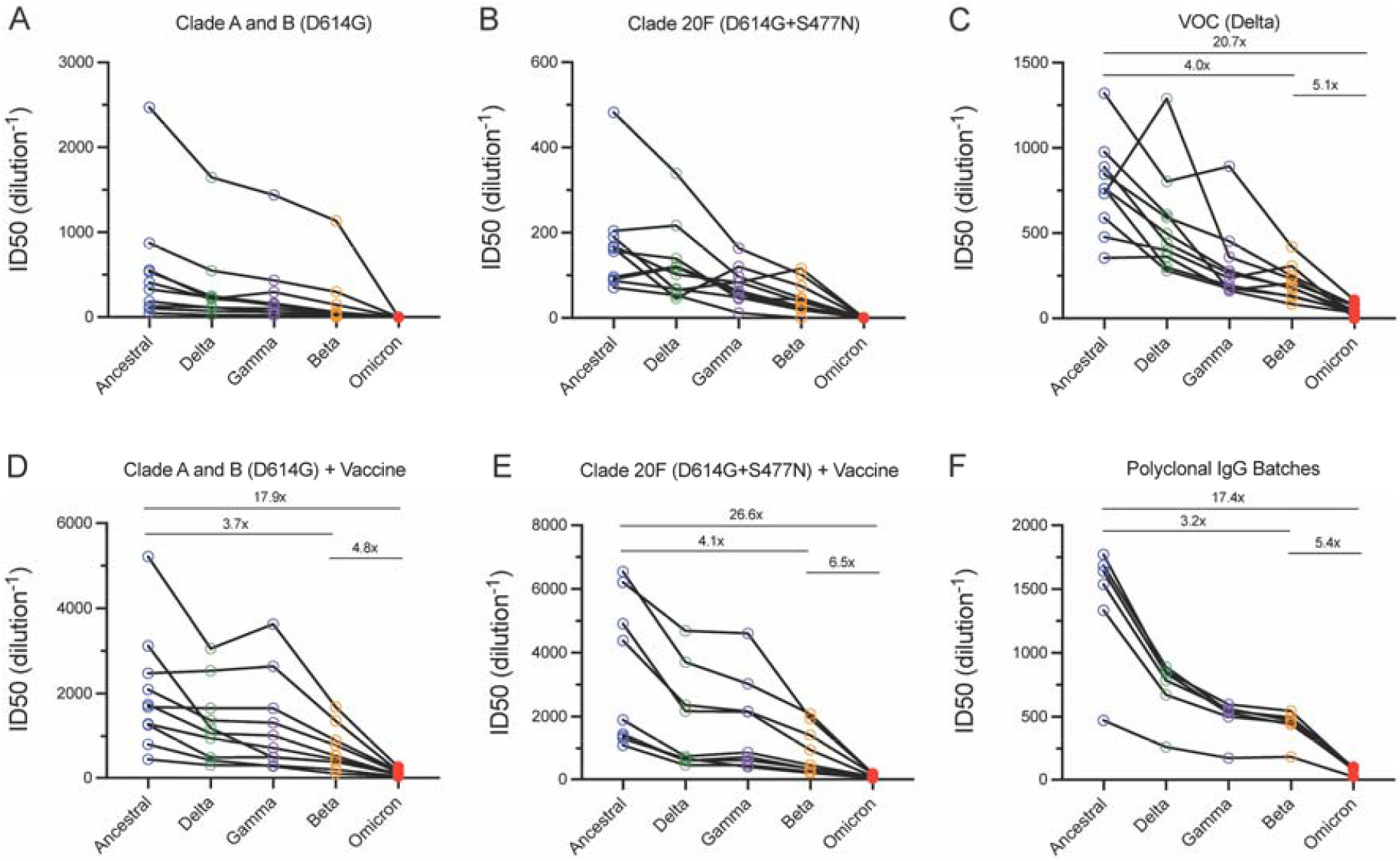
Humoral neutralisation of live SARS-CoV-2 variants in convalescent and vaccinated donors and concentrated human IgG plasma samples. Neutralisation assays were performed in a high-throughput format in HAT-24 cells using live virus isolates from the variants of concern; Delta (B.1.617.2), Gamma (P.1), Beta (B.1.351), Omicron (B.1.1.529) and the ancestral Wuhan-like virus with the original D614 background (A2.2) as a control. ID50 neutralisation titres presented for 5 live variants for convalescent donors from **(A)** Clade A and B (D614G) (first wave), **(B)** Clade 20F (D614G + S477N) (second wave) and **(C)** third (VOC Delta) wave. ID50 neutralisation titres presented for 5 live variants for vaccinated donors from **(D)** Clade A and B (D614G) (first wave) and **(E)** Clade 20F (D614G + S477N) (the second wave). **(F)** ID50 neutralisation titres across 5 live variants with concentrated human IgG from a mixture of vaccinated/convalescent and non-convalescent donors. **(C-F)** Fold change reductions in ID50 neutralisation titres compare the ancestral variant to Beta/Omicron and Beta to Omicron. Data in **(A-F)** is the mean of technical replicates in duplicate. *p<0.05, **p<0.01, ***p<0.001, ****p<0.0001 for Kruskal Wallis test with Dunn’s multiple comparison test.

## Supplementary Tables

Supplementary Table S1: Neutralisation assay on sera from vaccinated donors.

Supplementary Table S2: ADAPT participants and ID50s

Supplementary Table S3. SARS-CoV-2 variants used in this study.

| Variant | Lineage | Seq ID | GISAID ID |
| --- | --- | --- | --- |
| Ancestral | A.2.2 | nCoV_2715 | hCoV-19/Australia/NSW4715/2021 |
| Beta | B.1.351 | nCoV_2463 | hCoV-19/Australia/NSW4463/2021 |
| Gamma | P.1 | nCoV_2319 | hCoV-19/Australia/NSW4318/2021 |
| Delta | B.1.617.2 | nCoV_2605 | hCoV-19/Australia/NSW4605/2021 |
| Omicron | B.1.1.529 | RCoV_1933 | hCoV-19/Australia/NSW-RPAH-1933/2021 |

